# Influenza may facilitate the spread of SARS-CoV-2

**DOI:** 10.1101/2020.09.07.20189779

**Authors:** Matthieu Domenech de Cellès, Jean-Sébastien Casalegno, Bruno Lina, Lulla Opatowski

**Author notes:** **Corresponding author:** Dr. Matthieu Domenech de Cellès, Max Planck Institute for Infection Biology, Charitéplatz 1, Campus Charité Mitte, 10117 Berlin, Germany.

## Abstract

As in past pandemics, co-circulating pathogens may play a role in the epidemiology of coronavirus disease 2019 (COVID-19), caused by the novel severe acute respiratory syndrome coronavirus 2 (SARSCoV-2). Here we hypothesized that influenza interacted with SARS-CoV-2 during the early 2020 epidemic of COVID-19 in Europe. We developed a population-based model of SARS-CoV-2 transmission, combined with mortality incidence data in four European countries, to test a range of assumptions about the impact of influenza. We found consistent evidence for a 2–2.5-fold population-level increase in SARSCoV-2 transmission associated with influenza during the period of co-circulation. These results suggest the need to increase vaccination against influenza, not only to reduce the burden due to influenza viruses, but also to counteract their facilitatory impact on SARS-CoV-2.

## Main Text

The current pandemic of coronavirus disease 2019 (COVID-19), caused by the novel severe acute respiratory syndrome coronavirus 2 (SARS-CoV-2), has led to global alarm. Following the first case reports in December 2019 in Wuhan, China [1], SARS-CoV-2 rapidly spread across the globe and has resulted in approximately 19 million cases and 700,000 deaths worldwide, as of August 6, 2020 [2]. Because of the current lack of prophylactic or therapeutic treatments, the pandemic has caused the implementation of unprecedented control measures, which culminated in the lockdown of several billion people in over 100 countries during April–May 2020 [3]. Although a number of fixed (e.g., greater age, male sex) and chronic (e.g., hypertension, diabetes) risk factors of mortality have now been identified [4], the time-varying drivers of COVID-19 epidemiology remain poorly understood. Experience gained from past pandemics has highlighted the potentially large contribution of co-circulating pathogens to the burden of an emerging disease [5]. Despite the relevance for epidemic forecasting and for designing control strategies, the impact of co-circulating pathogens on SARSCoV-2 epidemiology has remained largely unexplored [6].

Respiratory viruses—including SARS-CoV-2 and other coronaviruses, rhinoviruses, influenza viruses, etc.—form a large class of viruses that cause seasonal infections of the respiratory tract in humans. Mounting evidence indicates that their epidemiologies are not independent, as a result of interaction mechanisms that may operate at different scales and that can be classified as either facilitatory or antagonistic [7, 8]. The interaction between the respiratory syncytial virus (RSV) and influenza may provide an example of antagonism. Indeed, experimental evidence in ferrets has shown that influenza viruses induce an antiviral state that transiently limits secondary infection with RSV [9], an effect postulated to explain the delayed epidemic of RSV during the 2009 influenza pandemic [10, 11]. Although such antagonistic interactions appear, to date, to be the most common among respiratory viruses [8], experimental evidence indicates that co-infections may have a facilitatory effect, for example by increasing viral growth [12]. Increased transmission of influenza during co-infection with other respiratory viruses was also proposed to explain the multiple waves during the 1918 influenza pandemic [13]. Interestingly, according to recent evidence a viral respiratory infection (in particular with influenza viruses) can up-regulate the expression of ACE2—the cognate receptor of SARS-CoV-2 in human cells—in the respiratory epithelium [14]. This suggests that respiratory viruses could affect the epidemiology of SARS-CoV-2. Here, we hypothesized that influenza—which peaked in February 2020 and therefore co-circulated during the early spread of COVID-19 in Europe (Fig. 1B)—interacted with SARS-CoV-2.

**Figure 1:**
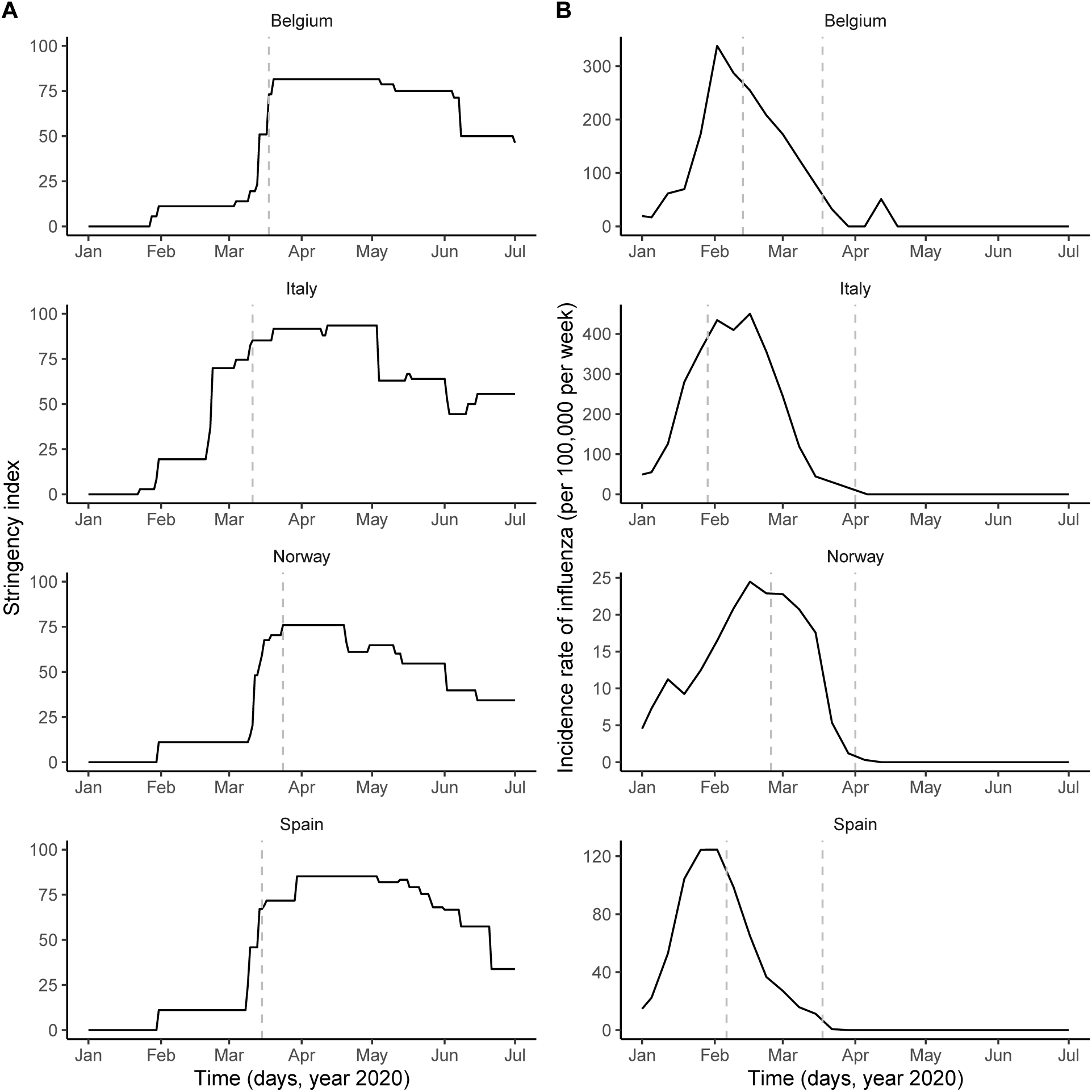
Potential drivers of SARS-CoV-2 transmission in Belgium, Italy, Norway, and Spain. A: time plot of the stringency index, a country-level aggregate measure of the number and of the strictness of non-pharmaceutical control measures implemented by governments. The vertical dashed line indicates the start of the nationwide lockdown [16]. B: time plot of influenza incidence, calculated as the product of the incidence of influenza-like illnesses and of the fraction of samples positive to any influenza virus (see also Fig. S1 for a time plot of the latter two variables). The vertical dashed lines delimitate the period of overlap between SARS-CoV-2 and influenza, defined as the period between the assumed start date of SARS-CoV-2 community transmission and 6 weeks after the epidemic peak of influenza [46]. In each country, the time series displayed were incorporated as covariates, which modulated the transmission rate of SARS-CoV-2 in our model (see Methods). In B, the *y*-axis values differ for each panel.

To test this hypothesis, we developed a stochastic, population-based model of SARS-CoV-2 transmission and of COVID-19 mortality [15, 16, 17]. Our model incorporated a realistic distribution of the generation time (mean 6.5 days, coefficient of variation 0.58, see Fig. S2 and Ref. [18]) and of the time from symptom onset to death (mean 17.8 days, coefficient of variation 0.45 [19]), under the assumption that 1% of all infections resulted in death [16]. A novel feature of our model was the inclusion of the stringency index, an aggregate measure of the number and of the strictness of non-pharmaceutical control measures (e.g., restrictions on travel, school or work closure, lockdown) implemented by governments in response to COVID-19 (Fig. 1A and ref. [3]). Specifically, we mapped the stringency index into a relative reduction in SARSCoV-2 transmission using a simple linear scaling function, whose slope represented the impact of control measures (see Methods). To assess the potential impact of influenza, we also incorporated renormalized time series of influenza incidence as drivers of SARS-CoV-2 transmission into our model. Using state-of-the-art statistical inference methods [20, 21], we systematically evaluated how different hypotheses about the impact of influenza explained the observed dynamics of COVID-19 mortality.

Based on influenza data availability from the World Health Organization (WHO, Figs. 1B, S1 and ref. [22]), we conducted our analysis in four European countries (Belgium, Italy, Norway, and Spain), where COVID-19 mortality peaked in March–April 2020 (Refs. [23, 24, 25, 26] and Fig. 2B). The results were unequivocal: we found consistent evidence that, during the period of co-circulation, influenza was associated with an average 2–2.5-fold population-level increase in SARS-CoV-2 transmission (Table 1). After controlling for the impact of influenza, our estimates of the basic reproduction number (*R*_0_) ranged from 2 (Italy and Spain) to 3.3 (Belgium). Although the increased transmission associated with influenza early during the SARS-CoV-2 epidemic explained the data significantly better (Table 1), a model without influenza led to higher *R*_0_ estimates (range 2.5–5, Fig. 2A), consistent with those of a previous study [16]. Also in line with [16], we found consistent evidence for a marked impact of non-pharmaceutical control measures (Table 1), which were associated with a decrease in SARS-CoV-2 transmission below the reproduction threshold from mid-March to early June 2020 (Fig. 2A). Visual inspection of simulations suggested that our model correctly captured the dynamics of COVID-19 mortality in every country (Fig. 2B). A more detailed model–data comparison of summary statistics confirmed that our model accurately predicted the peak time, the peak number and the total number of deaths, and the death growth exponent [27], except in Spain where the latter statistic was systematically under-estimated (Table S5). Our model-based estimates of the total proportion of individuals infected with SARS-CoV-2 (as of 4 May 2020, Table 1) were also comparable with those of a previous modeling study [16] and of a seroprevalence study conducted in early May in Spain [28]. Hence, our model appeared to precisely recapitulate the epidemiology of SARS-CoV-2 morbidity and mortality over a period of *∼*4 months.

**Figure 2:**
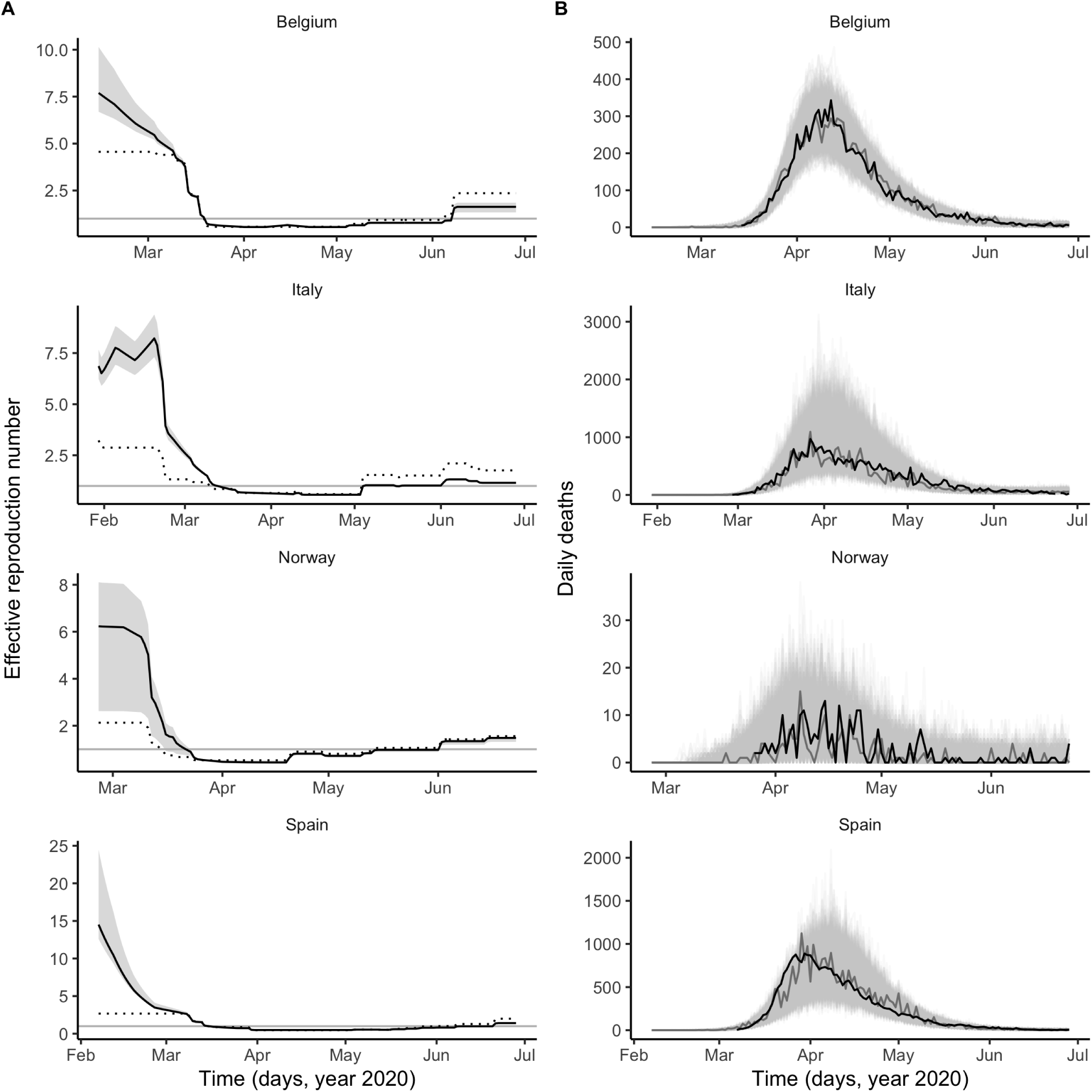
Dynamics of SARS-CoV-2 transmission and of COVID-19 mortality in Belgium, Italy, Norway, and Spain. A: time plot of the estimated effective reproductive number (*R_e_*). In each panel, the black line represents the maximum likelihood estimate and the grey ribbon the 95% confidence interval (calculated based on the likelihood profile of the influenza impact parameter, cf. Table 1) in each country. The dotted black line represents the effective reproduction number estimated from a model without influenza(i.e., with the influenza impact parameter fixed to 0 and the other parameters estimated from the data). The horizontal grey line is at *R_e_* = 1. B: time plot of the simulated and observed numbers of daily deaths caused by SARS-CoV-2. In each panel, the light grey lines represent 1,000 model simulations, with one simulation highlighted in dark grey; the black line represents the actual death counts. In A and B, the *x*-axis and the *y*-axis values differ for each panel.

**Table 1:** Model parameter estimates in Belgium, Italy, Norway, and Spain. For the proportion infected as of May 4, the numbers between parentheses represent a 95% prediction interval, based on 1,000 simulations at the maximum likelihood estimate. For the other parameters, they represent an approximate 95% confidence interval, calculated using either the profile likelihood [47] (parameter *e^βF^*) or a parametric bootstrap (other parameters). SE: standard error, calculated using 5 replicate particle filters, each with 20,000 particles, at the maximum likelihood estimate.

| Quantity | Belgium | Italy | Norway | Spain |
| --- | --- | --- | --- | --- |
| Study period (year 2020) | 13 Feb–28 Jun | 29 Jan–28 Jun | 25 Feb–28 Jun | 06 Feb–28 Jun |
| Log-likelihood (SE) | −383.2 (<0.1) | −669.1 (<0.1) | −160.8 (<0.1) | −558.8 (0.1) |
| Basic reproduction number ($R_0$) | 3.3<br>(2.0, 4.0) | 2.0<br>(2.0, 2.3) | 2.3<br>(2.0, 2.7) | 2.0<br>(2.0, 2.6) |
| Impact of control measures ($b$) | 1.02<br>(0.90, 1.06) | 0.76<br>(0.75, 0.81) | 1.07<br>(0.94, 1.09) | 0.89<br>(0.88, 0.96) |
| Average relative variation in SARS-CoV-2 transmission rate associated with influenza ($e^{\beta_F}$) | 1.9<br>(1.5, 2.8) | 2.4<br>(2.2, 2.7) | 2.1<br>(1.2, 2.8) | 2.5<br>(2.1, 3.1) |
| Initial number exposed to SARS-CoV-2 ($E_1(0)$) | 60<br>(10, 100) | 40<br>(15, 85) | 50<br>(80, 1550) | 130<br>(45, 540) |
| Proportion infected, as of 4 May 2020 (%) | 8.1<br>(6.1, 10.5) | 6.1<br>(4.2, 8.3) | 0.4<br>(0.3, 0.6) | 5.5<br>(4.4, 6.6) |

To verify the robustness of our results, we conducted three additional analyses (see Supplementary Results). First, we estimated an extended model in which influenza was allowed to modulate the lethality, in addition to the transmission, of SARS-CoV-2 (Table S2). Compared with the base model, we found little support for such a model in Belgium, Italy, and Norway (log-likelihood difference [∆log *L*] in the range 0.0–0.4, likelihood ratio test P-value in the range 0.37–1.00). In Spain, however, the parameter estimates suggested that influenza was associated with both increased transmission and lethality, but the statistical evidence was weak (∆log *L* = 3.2, *P* = 0.01). Second, we estimated another model, in which the reduction of SARS-CoV-2 transmission was allowed to scale non-linearly with the stringency index (Table S3). We found little statistical evidence that such a model outperformed the simple linear scaling function in Belgium, Norway, and Spain (∆log *L ∈* [0.1, 1.5], *P ∈* [0.08, 0.65]). In Italy, however, we found strong evidence (∆log *L* = 21.3, *P* < 10^−3^) for super-linear scaling at low values of the stringency index (Fig. S3), although our estimate of the impact of influenza was unchanged. This result may reflect the impact of the early lockdown in part of Lombardy, which preceded the one in Italy as a whole by a few weeks [29]. Third, to take into account potential spatial effects in the transmission dynamics of SARS-CoV-2 in Italy and in Spain [29, 28], we relaxed the assumption of homogeneous mixing (Table S4 and Refs. [30, 31]). In Italy (∆log *L* = 26.1, *P* < 10^−3^), but not in Spain (∆log *L* = 0.0, *P* = 1), we found evidence that the force of infection scaled sub-linearly with SARS-CoV-2 infection prevalence. This result may be explained by the progressive spread of SARS-CoV-2 from northern to southern regions of Italy [29]. Crucially, however, our estimate of the impact of influenza was unchanged in both countries. In addition, we found that our parameter estimates varied little when testing alternative hypotheses about the fixed value of the average generation time, of the onset-to-death time, and of the infection fatality ratio (Table S6). In sum, our main result about the impact of influenza remained robust to a variety of alternative assumptions regarding the epidemiological traits of SARS-CoV-2 and the modeled impact of control measures.

Our model makes at least two testable predictions. First, even though our results did not allow to distinguish between higher transmissibility or higher susceptibility in individuals co-infected with influenza and SARS-CoV-2, previous experimental work suggests that the latter mechanism may operate, as a result of up-regulation of the ACE2 receptor caused by influenza infection [14]. Hence, we predict that a recent influenza infection should be an independent risk factor for subsequent SARS-CoV-2 infection. Estimates of the frequency of co-detection of influenza and SARS-CoV-2 by polymerase chain reaction (PCR) testing in nasopharyngeal swabs were highly variable in previous studies (range 0–60% [6, 32]). Although the marked seasonality of influenza in temperate regions likely explains in part the low frequency found in some studies [32], we propose that differences in the natural history of influenza and SARS-CoV-2 infections also lead to a systematic under-estimation of co-infection. Specifically, because the incubation period of SARS-CoV-2 infection (estimated to average 5.7 days [33]) exceeds that of influenza (A, 1.4 days or B, 0.6 days [34]), it is likely that, by the time SARS-CoV-2 infection becomes detectable, influenza no longer is. To make that statement more precise, we calculated the probability of detectability of a co-infection, with influenza first then SARS-CoV-2 (Table S7). Assuming that influenza is detectable by PCR up to 4–5 days after [35], and SARS-CoV-2 from 2–4 days before [36], symptom onset, we find that a large, 30–50% of co-infections may not be detectable at all. These results may help explain the low frequency of co-detection found in some studies [37], and suggest that the time window of co-detectability may be too short to adequately infer the association between influenza and SARS-CoV-2 using PCR testing. Serological studies comparing the prevalence of antibodies against influenza in SARS-CoV-2 cases and non-cases may therefore be required to test the prediction that influenza is a risk factor for SARS-CoV-2 infection. Second, we predict that individuals vaccinated against influenza should be at lower risk of SARS-CoV-2 infection than those unvaccinated. The findings of a negative association between influenza vaccine coverage and COVID-19 mortality in ecological studies (in Italy [38] and in other countries [39]) and of a lower risk of SARS-CoV-2 infection in influenza vaccinees in a US prospective study [40] are consistent with our prediction, but further epidemiological investigations are needed. Importantly, our results can explain these findings as the direct effect of influenza vaccines on influenza infection, instead of indirect effects on non-influenza pathogens (e.g., as a result of trained immunity) [41].

Our study has a number of important limitations. First, for simplicity and as in other studies [15, 17, 16], our model was not age-structured, even though many aspects of COVID-19 and of influenza epidemiology— like disease severity and lethality—vary markedly with age [19]. The susceptibility to SARS-CoV-2 infection was also found to increase with age [42], a finding potentially explained by lower baseline expression of the ACE2 receptor in children [43]. Another testable prediction of our model, therefore, is that influenza should be associated with a transient increase in susceptibility to SARS-CoV-2 infection, commensurate with the variations of influenza incidence over age. Second, we modeled the impact of non-pharmaceutical control measures using a simple, linear function scaling the stringency index to the reduction of SARS-CoV-2 transmission. Even though this simple hypothesis provided a more parsimonious fit (except in Italy), that result may be specific to Europe, where control measures gradually increased in number and in intensity (Fig. 1A). In general, the association is likely non-linear (e.g., if a high-impact intervention like a lockdown is implemented early on), and we therefore recommend testing a variety of scaling functions. Third, we did not specifically model fully asymptomatic cases, which may represent a large fraction of SARS-CoV-2 infections [17]. The omission of asymptomatic infections may lead to biased *R*_0_ estimates if their duration significantly differs from that of symptomatic infections [44]. A previous study, however, estimated that the duration of both types of infection is comparable [17], such that our estimates should be robust in more complex model structures. Finally, we assessed only the impact of influenza, because of its high prevalence and period of overlap with SARS-CoV-2 in early 2020 in Europe and of the availability of high-quality data [22]. Nevertheless, other respiratory viruses, like RSV and rhinoviruses, may also interact with SARSCoV-2 and should be considered [14], especially if SARS-CoV-2 continues to spread widely during new seasons in temperate regions of the Northern hemisphere.

In conclusion, our results suggest that influenza virus infection increases the transmission of SARS-CoV-2 and facilitated its spread during the early 2020 epidemic of COVID-19 in Europe. Hence, an increase in the uptake of influenza vaccines may be called for, not only to reduce hospitalizations due to influenza infections [32, 45], but also to reduce their downstream impact on SARS-CoV-2 transmission and on COVID-19 mortality. More generally, taking into account the microbial environment of SARS-CoV-2 may be essential, not only to better understand its epidemiology, but also to enhance current and future infection control strategies.

## Data Availability

All data and R codes will be made available via a Dryad digital repository and are available upon request.

## Acknowledgements

We thank Arturo Zychlinsky and Klaus Osterrieder for helpful comments on the manuscript. Computations underlying the present analysis were performed at the Max Planck Computing and Data Facility (MPCDF)

## Authors contributions

MDdC conceived of the study design and performed the analysis. JSC and BL provided content expertise. LO conceived of the study design and oversaw the analysis.

## Data and materials availability

All data and R codes will be made available via a Dryad digital repository and are available upon request of the reviewers.

## Supplementary Materials

### S1 Materials and Methods

#### Data

##### Stringency index data

Country-level time series of the stringency index were available from the Oxford COVID-19 Government Response Tracker, developed at the University of Oxford and described elsewhere [3]. Briefly, the stringency index provides an aggregate measure of the number and of the strictness of non-pharmaceutical control measures implemented by governments in response to the COVID-19 epidemic. The stringency index is defined as the average of 9 normalized ordinal variables, which quantify the strength (e.g., recommended or required) and the scope (e.g., targeted or general) of closure and containment measures (8 variables) and of health measures (1 variable). The resulting index allows to quantify the strength of control measures in a systematic way, on a scale ranging from 0 (no interventions) to 100 (maximum number and maximal intensity of control measures). Of note, however, the stringency index does not quantify the impact of control measures, which likely varied across countries [16]. In formulating our model, we therefore modeled the relationship between the stringency index and the relative reduction in SARS-CoV-2 transmission using a non-decreasing function, whose parameters represented the impact of control measures and were estimated from the data.

##### Influenza incidence data

Virological data on the weekly numbers of samples tested and of samples positive to any influenza virus were available from the FluNet database, compiled by the WHO (Fig. S1A). Parallel syndromic data on the weekly incidence rate of influenza-like illnesses (ILI) were available from the FluID database, also compiled by the WHO (Fig. S1B). Those data were deemed high-quality and used in a previous study on influenza forecasting in the countries considered here [22]. The weekly incidence rate of influenza was then calculated as the product of ILI incidence and of the fraction of samples positive to any influenza virus (Fig. 1B). Because the magnitude of influenza incidence thus calculated varied markedly across countries (e.g., as a result of different surveillance systems and case definitions), we rescaled each time series by its average during the period of co-circulation of influenza and SARS-CoV-2 (Fig. 1B). The resulting time series was therefore dimensionless and equalled 1 when influenza equalled its average value during that period.

**Figure S1:**
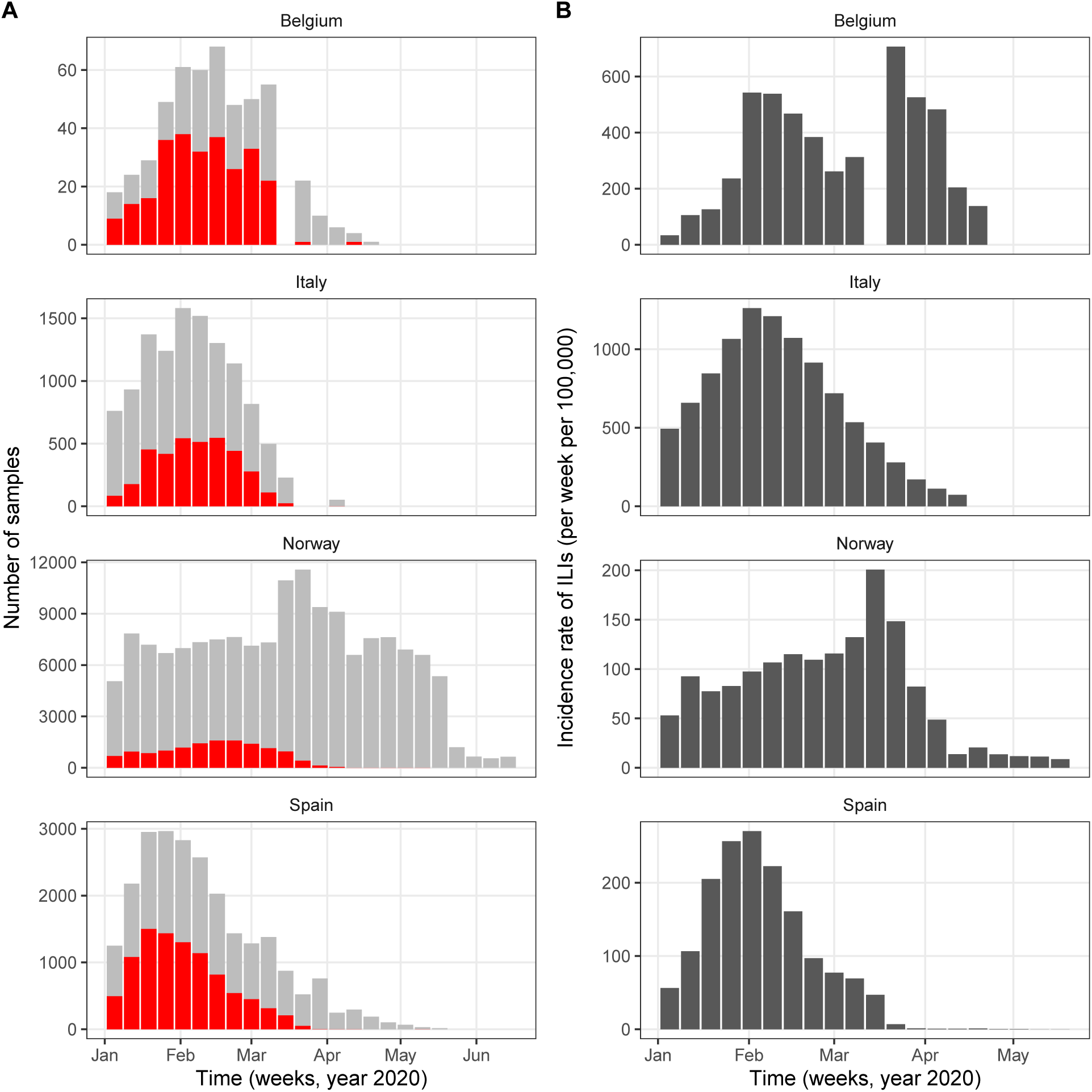
Syndromic ILI data (A) and virological influenza data (B) in Belgium, Italy, Norway, and Spain. In A, the red bars represent the numbers of samples positive to any influenza virus and the grey bars those negative.

##### COVID-19 mortality data

Data on the daily number of deaths caused by SARS-CoV-2 (counted by date of death) were available from national public health public institutes, in Belgium (Sciensano [23]), in Italy (*Dipartimento della Protezione Civile* [24]), and in Spain (*Instituto de Salud Carlos III*, official data with historical corrections compiled by the media DATADISTA [26]). In Norway, the data were available from the worldwide database compiled by the European Center for Disease Control and Prevention [25]. Following a previous study [16], and to avoid a possible bias caused by the dominance of deaths due to non-locally acquired infections early in the epidemic, we included observed deaths from the date after which the cumulative observed death count exceeded 10. Data points before that date were treated as missing and were assigned a conditional log-likelihood of 0, such that they did not contribute to the overall log-likelihood. The data were not further pre-processed, except in Italy, where a negative death count was reported on 24 June 2020 and was treated as missing and also assigned a log-likelihood of 0.

#### Transmission model

##### Model formulation

We formulated a variant of the standard Susceptible–Exposed–Infected–Recovered transmission model [48], using the method of stages to allow for a realistic distribution of the latent, infectious, and onset-to-death periods [49, 50]. Specifically, we assumed that the latent and infectious periods were Erlang-distributed with shape parameter 2 and mean 1*/σ* = 4 days and 1*/γ* = 5 days, respectively [17]. The resulting generation time *T_g_* (i.e., the time from infection of a primary case to transmission to a secondary case) had a mean of 6.5 days and a coefficient of variation of 0.58 (see Fig. S2 for the full distribution and the details of the calculation), consistent with empirical observations and with the values fixed in a previous modeling study [18, 16]. To model the impact of the gradual implementation of non-pharmaceutical control measures (e.g., border closure, school closure, lockdown), we mapped the stringency index (denoted by *s*(*t*)) to the time-varying relative reduction in transmission of SARS-CoV-2 (denoted by *r_β_*(*t*)). Specifically, we used the following simple linear scaling function, with saturation:

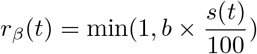

Here the parameter *b* quantifies how fast the transmission rate of SARS-CoV-2 decreases as the stringency index increases. Hence, this parameter can be interpreted as a measure of the impact of non-pharmaceutical control measures on SARS-CoV-2 transmission. The deterministic variant of the model was represented by the following set of differential equations:

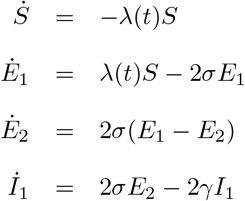

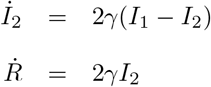

The force of infection (that is, the per capita rate at which susceptible individuals contract infection [48]), *λ*(*t*), was modeled as:

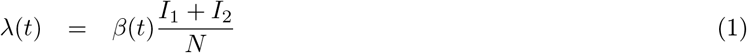

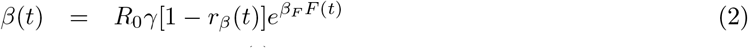

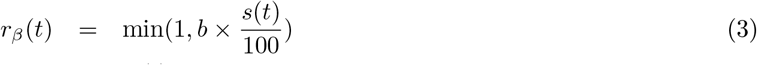

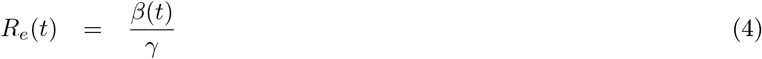

where *R*_0_ represents the basic reproduction number of SARS-CoV-2, *R_e_*(*t*) the time-varying effective reproduction number, *N* the population size (assumed constant during the study period), and *F* (*t*) the renormalized time series of influenza incidence, incorporated as a covariate into the model (Fig. 1). With this formulation, the parameter *β_F_* quantifies the impact of influenza on SARS-CoV-2 transmission: *β_F_* > 0 if influenza increases transmission, *β_F_* < 0 if influenza decreases transmission, and *β_F_* = 0 if influenza has no impact on transmission (null hypothesis). More specifically, the average incidence of influenza during the period of co-circulation with SARS-CoV-2 corresponds to *F* (*t*) = 1, such that *e^βF^* represents the average relative variation of SARS-CoV-2 transmission associated with influenza (null hypothesis: *e^βF^* = 1). In the main text and in the following text, we report the estimates of *e^βF^* to facilitate the interpretation of the impact of influenza. In writing equation (2), we implicitly assume that the impact of influenza on SARS-CoV-2 transmission, if any, is short-lived and does not extend long after influenza infection.

Finally, we incorporated an observation model that related the dynamics of SARS-CoV-2 infection to that of COVID-19 mortality, taking into account the fact that only a fraction of infections results in death and that, among those, death occurs some time after symptom onset [19, 33, 16]. We assumed an average duration of pre-symptomatic of 2.5 days, resulting in an average incubation duration of 6.5 days, in broad agreement with previous empirical studies [36, 33]. Hence, individuals in the first infected state (*I*_1_) were considered pre-symptomatic, and the onset of symptoms was assumed to coincide with the transition from *I*_1_ to *I*_2_. The onset-to-death time was then assumed to be Erlang distributed with shape parameter 5 and mean 1*/κ* = 17.8 days (coefficient of variation of 0.45), the value estimated in a previous epidemiological study [19]. In sensitivity analysis, we also tested a mean onset-to-death time of 1*/κ* = 13 days, the lower bound estimated in a meta-analysis [33]. According to previous studies, the infection fatality ratio (IFR) typically ranges from 0.5% to 1% [51, 19]. We fixed the IFR to *µ* = 0.01 in the base model, but we considered an alternative value of 0.005 in a sensitivity analysis. Given those assumptions, the observation model was modeled by the following set of ordinary differential equations:

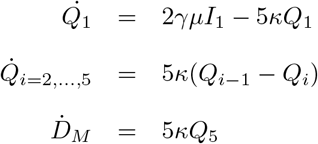

Here *D_M_* is the simulated number of daily deaths, modeled as an accumulator variable and reset to 0 at the end of each day. The observed number of daily deaths, *D_O_*, was modeled using a negative binomial distribution with mean *D_M_* and over-dispersion 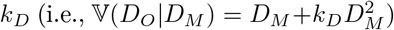, a standard distribution used in a number of previous modeling studies [16, 52].

As in Flaxman et al. [16], simulations were started 30 days before the date from which the cumulative observed death count first exceeded 10. At that date, we assumed that *E*_1_(0) individuals had been exposed to SARS-CoV-2; other individuals were assumed susceptible to infection (i.e., *S*(0) = *N* − *E*_1_(0)), and all other compartments were initialized to 0.

The stochastic variant of the model was implemented as a continuous-time Markov process approximated via a multinomial modification of the *τ*-leap algorithm [31], with a fixed time step of 10^−1^ day.

**Figure S2:**
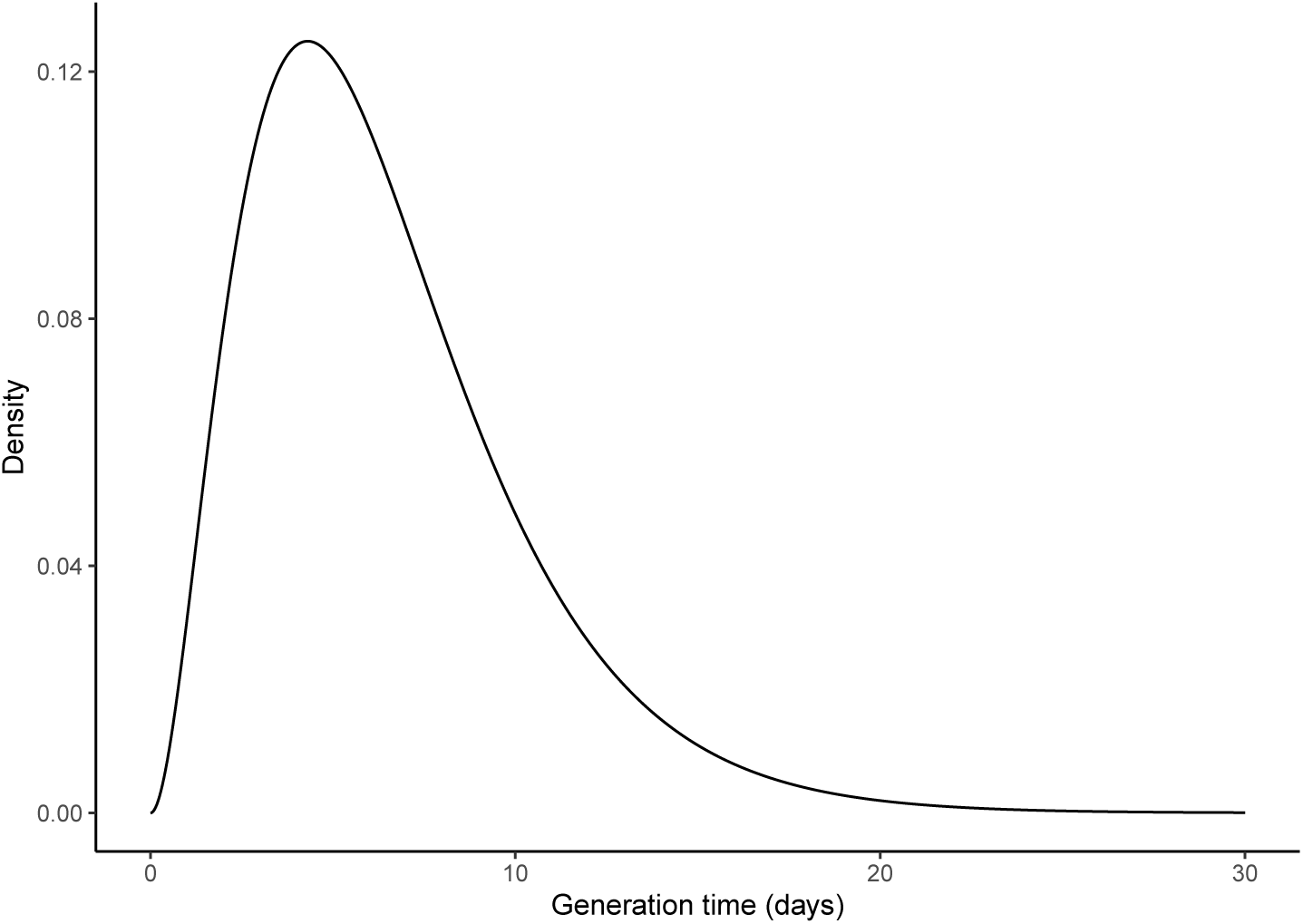
Model-based distribution of the generation time. For our model, the density function of the generation time is given by [53, 54]: 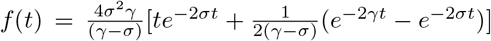. The resulting distribution is bell-shaped, with mean 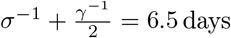 and coefficient of variation 0.58.

##### Model estimation

The following five parameters were estimated from the data:

1. The basic reproduction number, *R*_0_. According to a previous meta-analysis [55], this parameter was searched in the interval 2–10.
2. The impact of non-pharmaceutical control measures, *b*. The lower bound of the search interval of this parameter was fixed to 0.5, such that the maximal value of the stringency index (*s* = 100) corresponded to a minimal reduction of SARS-CoV-2 transmission of 50% [16].
3. The impact of influenza on SARS-CoV-2 transmission, *β_F_* (search interval: ℝ).
4. The initial number of individuals exposed to SARS-CoV-2, *E*_1_(0) (search interval: 0–10^4^).
5. The over-dispersion in death reporting, *k_D_* (search interval: ℝ^+^).

A summary list of fixed and estimated model parameters is presented in Table S1

All parameters were transformed to be estimated on an unconstrained scale, using a log transformation for positive parameters and the extended logistic function 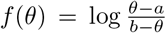 for parameters constrained in the interval [*a, b*]. The maximum iterated filtering algorithm (MIF2 [20]), implemented in the R package pomp [21, 56], was used to estimate model parameters. The estimation was completed in several steps, starting with trajectory matching to identify good starting parameters for MIF2, followed by 100 independent runs of MIF2 to locate the maximum likelihood estimate (MLE). The profile likelihood was calculated to verify the convergence of MIF2 and to derive an approximate 95% confidence interval for the parameter *β_F_* [47]. For the other parameters, a parametric bootstrap was used to calculate approximate 95% confidence intervals, by re-estimating the parameters for each of 200 synthetic datasets simulated at the MLE [57, 58].

**Table S1:**
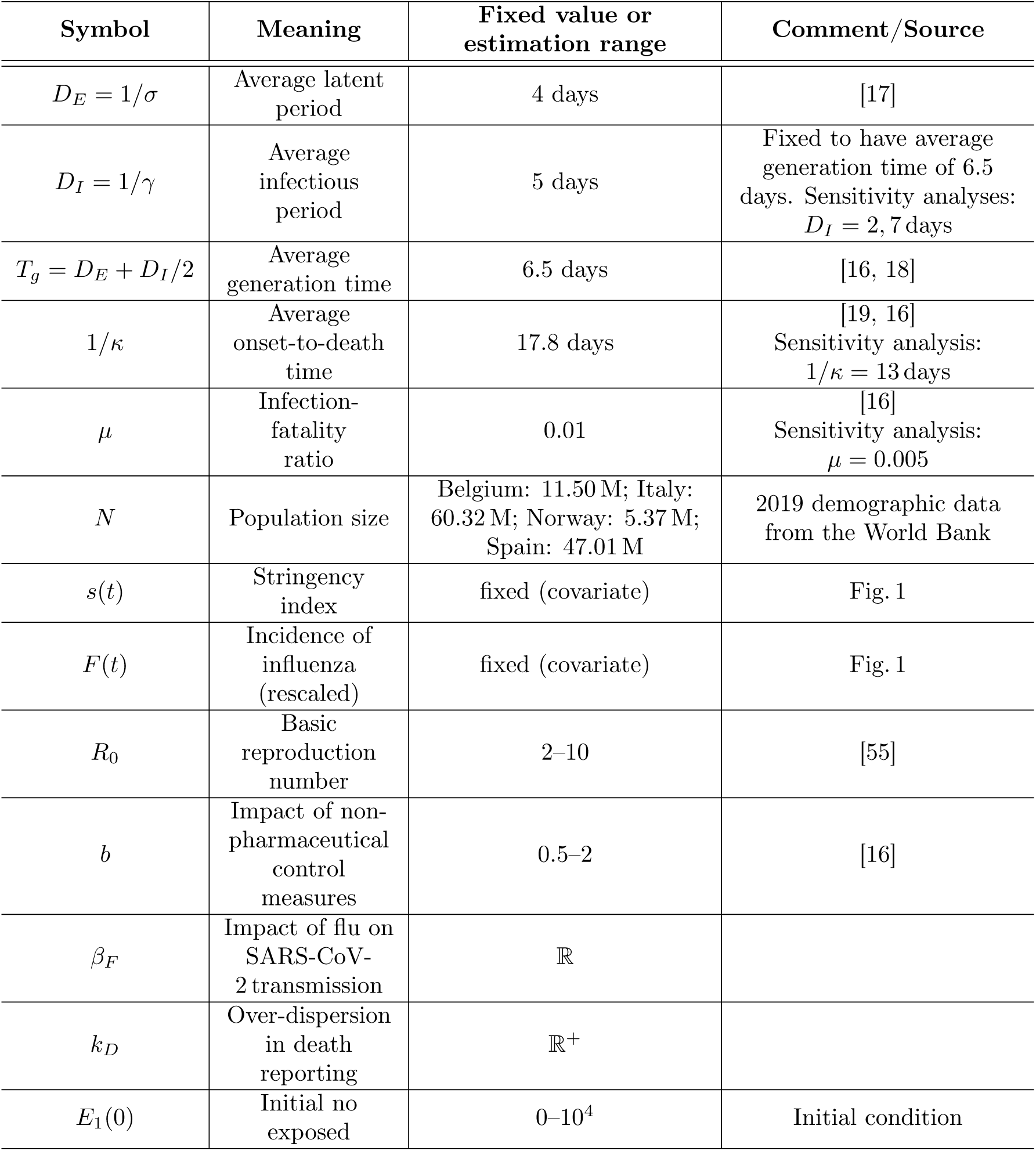
List of model parameters.

| Symbol | Meaning | Fixed value or estimation range | Comment/Source |
| --- | --- | --- | --- |
| $D_E = 1/\sigma$ | Average latent period | 4 days | [17] |
| $D_I = 1/\gamma$ | Average infectious period | 5 days | Fixed to have average generation time of 6.5 days. Sensitivity analyses: $D_I = 2, 7$ days |
| $T_g = D_E + D_I/2$ | Average generation time | 6.5 days | [16, 18] |
| $1/\kappa$ | Average onset-to-death time | 17.8 days | [19, 16]<br>Sensitivity analysis: $1/\kappa = 13$ days |
| $\mu$ | Infection-fatality ratio | 0.01 | [16]<br>Sensitivity analysis: $\mu = 0.005$ |
| $N$ | Population size | Belgium: 11.50 M; Italy: 60.32 M; Norway: 5.37 M; Spain: 47.01 M | 2019 demographic data from the World Bank |
| $s(t)$ | Stringency index | fixed (covariate) | Fig. 1 |
| $F(t)$ | Incidence of influenza (rescaled) | fixed (covariate) | Fig. 1 |
| $R_0$ | Basic reproduction number | 2–10 | [55] |
| $b$ | Impact of non-pharmaceutical control measures | 0.5–2 | [16] |
| $\beta_F$ | Impact of flu on SARS-CoV-2 transmission | $\mathbb{R}$ | |
| $k_D$ | Over-dispersion in death reporting | $\mathbb{R}^+$ | |
| $E_1(0)$ | Initial no exposed | 0–$10^4$ | Initial condition |

### S2 Supplementary results

#### Model with impact of influenza on both transmission and lethality

To test the hypothesis that influenza increases COVID-19 lethality, in addition to SARS-CoV-2 transmission, we extended our base model so that influenza was also allowed to modulate the IFR. Specifically, we replaced the constant IFR *µ* by the time-varying parameter:

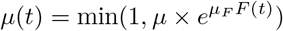

where *µ_F_ ∈* ℝ quantifies the impact of influenza on COVID-19 mortality and was estimated from the data. The results are presented in Table S2 and are discussed in the main text.

**Table S2:** Point parameter estimates of an extended model with impact of influenza on both transmission and mortality. ^*∗*^Log-likelihood difference (P-value, from a log-likelihood ratio test) with the base model presented in Table 1.^*†*^Parameter not identified.

| Quantity | Belgium | Italy | Norway | Spain |
| --- | --- | --- | --- | --- |
| $\log L$ (SE) | -383.2 (<0.1) | -668.7 (<0.1) | -160.8 (<0.1) | -555.6 (0.3) |
| $\Delta \log L$ (P)* | 0.0 (1.00) | 0.4 (0.37) | 0.0 (1.00) | 3.2 (0.01) |
| $R_0$ | 3.3 | 2.0 | 3.3 | 2.1 |
| $b$ | 1.02 | 0.77 | 0.90 | 0.90 |
| $e^{\beta_F}$ | 1.8 | 2.1 | 0.6 | 3.2 |
| $e^{\mu_F}$ | 0.9 | 0.6 | NA<sup>†</sup> | 1.83 |
| $k_D$ | $8.4 \times 10^{-4}$ | $1.1 \times 10^{-2}$ | $2.0 \times 10^{-1}$ | $7.2 \times 10^{-2}$ |
| $E_1(0)$ | 90 | 260 | 20 | 10 |

#### Model with non-linear function mapping the stringency index to the relative reduction in transmission

Although we assumed a simple linear scaling in our base model, it can also be hypothesized that the reduction of SARS-CoV-2 transmission scales non-linearly with the stringency index. For example, super-linear scaling for low values of the stringency index may occur if a potentially high-impact intervention (e.g., lockdown) is implemented early on, such that a modest increase in the stringency index results in a marked decrease in SARS-CoV-2 transmission. Conversely, sub-linear scaling may also be plausible if potentially low-impact interventions (e.g., border closure) are implemented first. To test those hypotheses, we considered an alternative, non-linear scaling function of the form:

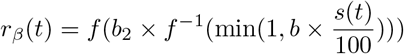

where *f*(*x*) = (1+*e^−x^*)^−1^ is the logistic function. Here the extra parameter *b*_2_ controls the slope at the origin, with *b*_2_ < 1 representing super-linear scaling at low values of the stringency index, and *b*_2_ > 1 super-linear scaling. For *b*_2_ = 1, 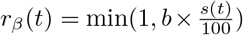), such that the base model with linear scaling is nested within this more general model. The corresponding parameter estimates are presented in Table S3; the scaling function estimated in Italy is also plotted in Fig. S3 and further discussed in the main text.

**Table S3:** Point parameter estimates of an extended model with a non-linear scaling function for the stringency index. ^*∗*^Log-likelihood difference (P-value from a log-likelihood ratio test) with the base model presented in Table 1.

| Quantity | Belgium | Italy | Norway | Spain |
| --- | --- | --- | --- | --- |
| $\log L$ (SE) | -383.1 (0.1) | -647.8 ($<0.1$) | -160.7 ($<0.1$) | -557.3 (0.1) |
| $\Delta \log L$ (P)* | 0.1 (0.65) | 21.3 ($< 10^{-3}$) | 0.1 (0.65) | 1.5 (0.08) |
| $R_0$ | 2.9 | 2.0 | 2.1 | 2.0 |
| $b$ | 0.87 | 0.99 | 0.91 | 0.94 |
| $b_2$ | 1.60 | 0.33 | 1.62 | 0.84 |
| $e^{\beta_F}$ | 1.9 | 2.5 | 2.1 | 2.9 |
| $k_D$ | $1.3 \times 10^{-3}$ | $6.6 \times 10^{-2}$ | $1.6 \times 10^{-1}$ | $7.9 \times 10^{-2}$ |
| $E_1(0)$ | 70 | 150 | 60 | 50 |

**Figure S3:**
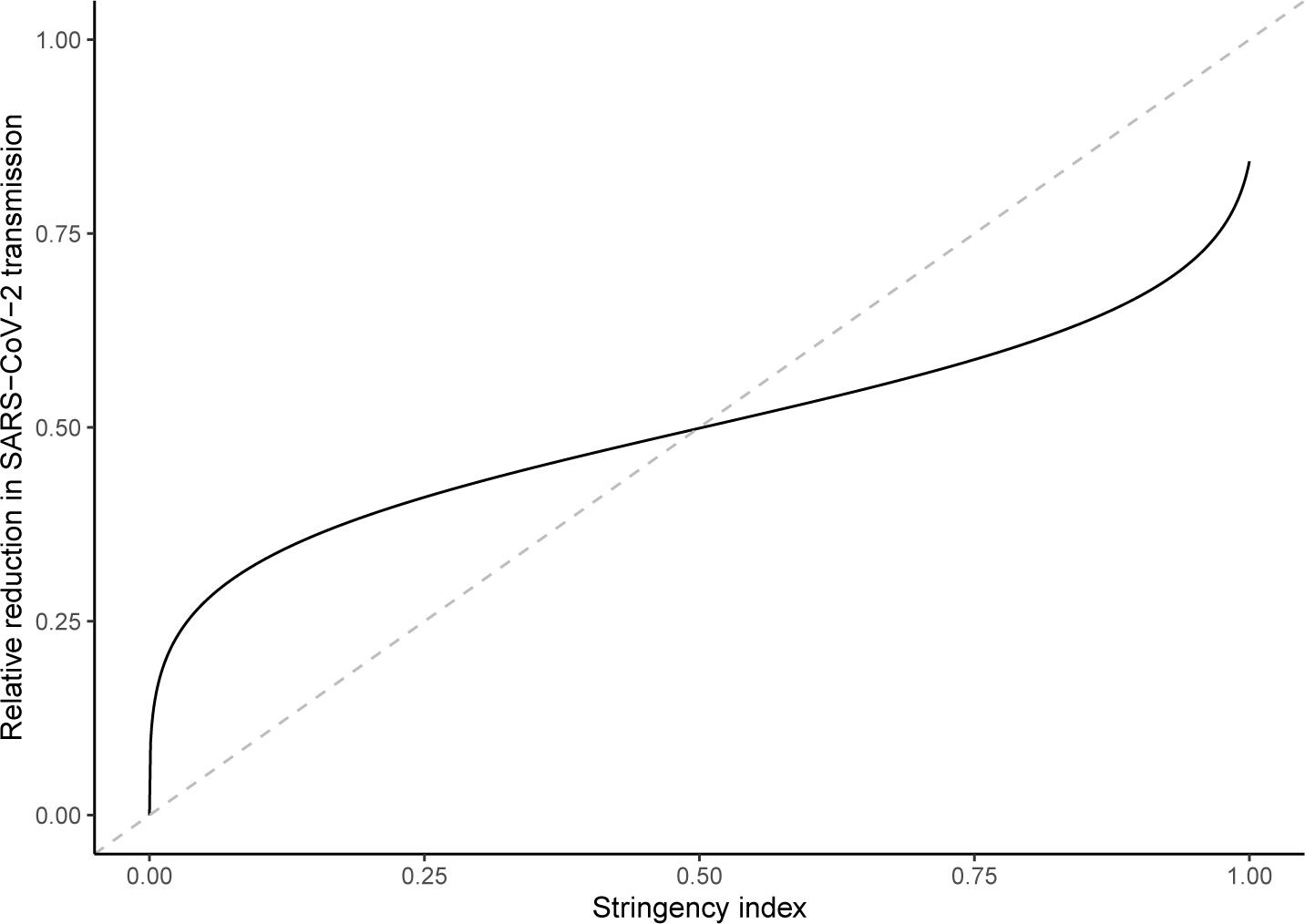
Estimated shape of the scaling function in Italy. The dashed grey line represents the identity line.

#### Model extension with inhomogeneous mixing

To model spatial effects in the dynamics of SARS-CoV-2 transmission in Italy and in Spain [29, 28], we implemented an extended model in which we relaxed the assumption of homogeneous mixing [30]. Here we assumed that the force of infection could scale non-linearly with SARS-CoV-2 infection prevalence [31]:

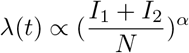

where the mixing coefficient *α ≥* 0 was estimated from the data. Given that 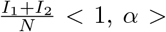 1 represents sub-linear scaling and *α* < 1 super-linear scaling. The results of those experiments are presented in Table S4 and further discussed in the main text.

**Table S4:** **Point parameter estimates of an extended model with** inhomogeneous mixing. *Loglikelihood difference (P-value) with the base model presented in Table 1.

| Quantity | Italy | Spain |
| --- | --- | --- |
| $\log L$ (SE) | -643.0 ($<0.1$) | -558.8 ($<0.1$) |
| $\Delta \log L$ (P)* | 26.1 ($< 10^{-3}$) | 0.0 (1.0) |
| $R_0$ | 2.1 | 2.1 |
| $b$ | 0.59 | 0.91 |
| $e^{\beta_F}$ | 2.5 | 2.4 |
| $k_D$ | $6.3 \times 10^{-2}$ | $8.0 \times 10^{-2}$ |
| $E_1(0)$ | 570 | 120 |
| $\alpha$ | 1.07 | 1.00 |

#### Model fit to data summary statistics

To evaluate the model fit in more detail, we examined the model– data agreement on a number of statistics that summarized important aspects of the mortality data—that is, probes [59, 21]). Specifically, we considered the following probes:

- Peak time (in days relative to the start of the study period).
- Peak daily number of deaths.
- Total number of deaths.
- Epidemic growth exponent. According to a previous study [27], we assumed that, until the peak time, the daily number of deaths grew algebraically, i.e., *D*(*t*) ∝ *t^α^*. We then estimated the growth exponent *α* using a log-log linear regression model.

The observed and simulated probe values are summarized in Table S5 and discussed in the main text.

**Table S5:**
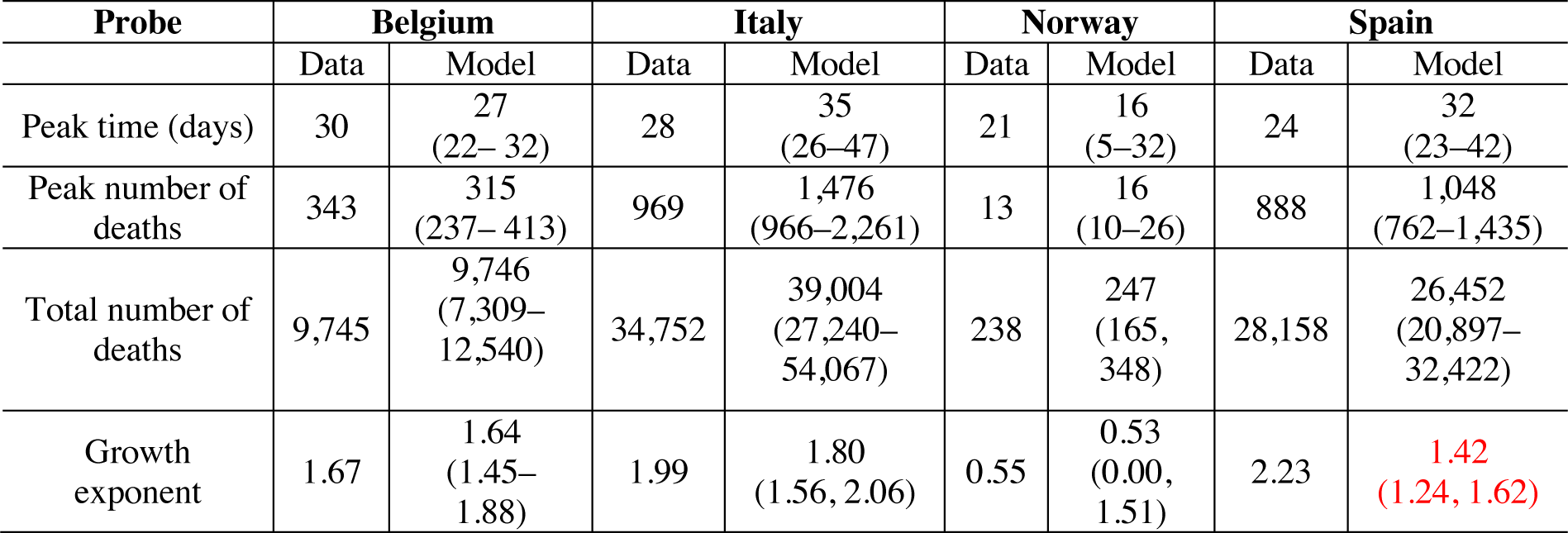
Model–data comparison on probes. The values highlighted in red indicate model-based probes that did not enclose the observed data probe value.

| Probe | Belgium |  | Italy |  | Norway |  | Spain |  |
| --- | --- | --- | --- | --- | --- | --- | --- | --- |
|  | Data | Model | Data | Model | Data | Model | Data | Model |
| Peak time (days) | 30 | 27<br>(22–32) | 28 | 35<br>(26–47) | 21 | 16<br>(5–32) | 24 | 32<br>(23–42) |
| Peak number of deaths | 343 | 315<br>(237–413) | 969 | 1,476<br>(966–2,261) | 13 | 16<br>(10–26) | 888 | 1,048<br>(762–1,435) |
| Total number of deaths | 9,745 | 9,746<br>(7,309–12,540) | 34,752 | 39,004<br>(27,240–54,067) | 238 | 247<br>(165, 348) | 28,158 | 26,452<br>(20,897–32,422) |
| Growth exponent | 1.67 | 1.64<br>(1.45–1.88) | 1.99 | 1.80<br>(1.56, 2.06) | 0.55 | 0.53<br>(0.00, 1.51) | 2.23 | 1.42<br>(1.24, 1.62) |

#### Sensitivity analyses

To verify the robustness of our results, we conducted a number of sensitivity analyses. Specifically, we modified the value of 3 fixed model parameters (infection fatality ratio, average onset-to-death time, and average generation time) and we repeated the estimations as before. As shown in Table S6, the estimate of the impact of influenza on SARS-CoV-2 transmission (*e^βF^*) remained consistently above 1 for all scenarios tested.

**Table S6:** Sensitivity analyses.

| Model | Belgium |  |  |  |  | Italy |  |  |  |  | Norway |  |  |  |  | Spain |  |  |  |  |
| --- | --- | --- | --- | --- | --- | --- | --- | --- | --- | --- | --- | --- | --- | --- | --- | --- | --- | --- | --- | --- |
| | $\log L$<br>(SE) | $R_0$ | $b_1$ | $e^{\beta_F}$ | $E_1(0)$ | $\log L$<br>(SE) | $R_0$ | $b_1$ | $e^{\beta_F}$ | $E_1(0)$ | $\log L$<br>(SE) | $R_0$ | $b$ | $e^{\beta_F}$ | $E_1(0)$ | $\log L$<br>(SE) | $R_0$ | $b$ | $e^{\beta_F}$ | $E_1(0)$ |
| Base model | −383.2<br>(<0.1) | 3.3 | 1.02 | 1.9 | 60 | −669.1<br>(<0.1) | 2.0 | 0.76 | 2.4 | 40 | −160.8<br>(<0.1) | 2.3 | 1.07 | 2.1 | 50 | −558.8<br>(0.1) | 2.0 | 0.89 | 2.5 | 130 |
| $E(T_g) = 5$ days | −384.4<br>(0.1) | 2.8 | 0.94 | 1.5 | 30 | −688.4 | 2.0 | 0.73 | 1.9 | 10 | −161.1<br>(<0.1) | 2.1 | 0.97 | 1.5 | 70 | −562.5<br>(0.1) | 2.0 | 0.84 | 1.5 | 340 |
| $E(T_g) = 7.5$ days | −382.3<br>(<0.1) | 3.5 | 1.06 | 2.4 | 50 | −662.9<br>(<0.1) | 2.0 | 0.79 | 2.8 | 40 | −160.7<br>(<0.1) | 2.5 | 1.12 | 2.5 | 60 | −556.4<br>(0.4) | 2.0 | 0.94 | 3.8 | 20 |
| $\mu = 0.005$ | −383.3<br>(<0.1) | 3.7 | 1.02 | 1.7 | 140 | −661.9<br>(<0.1) | 2.0 | 0.74 | 2.3 | 90 | −160.5<br>(<0.1) | 2.3 | 1.07 | 2.2 | 100 | −557.4<br>(0.1) | 2.0 | 0.88 | 2.7 | 110 |
| $\kappa^{-1} = 13$ days | −395.7<br>(0.1) | 2.0 | 0.88 | 7.6 | 1 | −692.8<br>(0.1) | 2.0 | 0.76 | 3.1 | 2 | −162.3<br>(0.1) | 2.1 | 1.00 | 2.8 | 15 | −538.8<br>(0.4) | 2.0 | 0.87 | 4.7 | 2 |

#### Probability that an influenza–SARS-CoV-2 co-infection is detectable

To calculate the probability that a co-infection with influenza then SARS-CoV-2 can be be detected, we ran a simulation study. Assuming that influenza infection occurred first, we first generated a sample of influenza incubation periods from a log-Normal distribution with median 1.4 days and dispersion 1.51, based on the results of a previous review [34]. We then generated a sample of detection periods, assuming that influenza could be shed (and therefore detected) up to 4–5 days after symptom onset [35]. Second, we generated a sample of SARS-CoV-2 infection times, uniformly between the time of infection and the end time of detectability of influenza. Finally, we generated a sample of SARS-CoV-2 incubation periods (from a Gamma distribution with mean 5.7 days [33] and coefficient of variation 0.86 [16]) and of SARS-CoV-2 detection start times, assuming that SARS-CoV-2 could be detected from 2 to 4 days before symptom onset [36]. In each simulation, we calculated the probability that co-detection can be detected as the fraction of the sample for which the maximal detection time of influenza exceeded the minimal detection time of SARS-CoV-2. The results are presented in Table S7 and discussed in the main text.

**Table S7:** Probability that an influenza–SARS-CoV-2 co-infection can be detected. The results are based on sample size of 10^5^; replicate simulations gave identical results, such that the estimates may be considered exact.

| Detection time of<br>influenza after<br>symptom onset | Detection time of<br>SARS-CoV-2 before<br>symptom onset | Probability of<br>co-detection |
| --- | --- | --- |
| 4 days | 2 days | 0.52 |
| 4 days | 4 days | 0.67 |
| 5 days | 2 days | 0.55 |
| 5 days | 4 days | 0.70 |

